# Metabolic health specific functional connectivity signatures in the human brain

**DOI:** 10.64898/2026.02.06.26345776

**Authors:** Kenneth Yuen, Isabel Arend

## Abstract

Obesity and metabolic dysfunction are among the strongest risk factors for poor brain and mental health, yet the neural mechanisms linking metabolism, brain, and behaviour remains poorly understood. Here, we provide the first evidence for two distinct large-scale brain network configurations—one associated with metabolic health and another with obesity— identified using resting-state fMRI data and metabolic phenotypes from a large community cohort (*N* = 564). While obesity was linked to enhanced coupling between subcortical reward and higher-order cortical networks, metabolic health was characterized by functional integration among default mode, salience, and frontoparietal control regions (metabolic health functional connectivity; MHFC). The MHFC network mediated the relationship between eating restraint and metabolic health, independent on individual’s body weight and metabolic status, and it was replicated with data from a different time point. Longitudinal analysis showed that change of MHFC strength predicted metabolic indicators over time, suggesting a role for this network as a potential marker of metabolic resilience. These findings reveal a neurobiological pathway through which executive and interoceptive regulatory systems contribute to metabolic health, offering new insights into the brain mechanisms linking eating behaviour, metabolism, and brain function.

## Introduction

Obesity is a major risk factor for cardiometabolic disease, neurodegeneration, and accelerated aging. Yet, the presence of metabolically healthy individuals with obesity—and, conversely, lean individuals with metabolic dysfunction—highlights a physiological heterogeneity that challenges traditional classifications and remains poorly understood^1^.

Obesity is a multifactorial condition with genetics, neurobiological, and psychosocial factors that interact, giving raise to different phenotypes. Functional connectivity reflects how anatomically distinct brain regions form coherent networks that support different functions. Neuroimaging research increasingly supports a model of obesity as a disorder of altered brain network dynamics^2-4^. Under task conditions, functional connectivity reveals how network dynamics relate to behavior. In obesity, task-evoked connectivity studies using reward- and food-cue paradigms show hyperconnectivity in reward-related regions, reflecting the heightened motivational value of palatable foods, and hypoconnectivity across control networks, indicating weakened self-regulation and inhibitory control^3^.

While task-based studies link context-dependent responses to food and reward cues with obesity, they do not capture obesity-specific neural organization. Studies examining functional connectivity under rest (i.e., resting-state functional connectivity; rsFC) reveal hypoconnectivity in the default mode (DMN) and executive control networks (ECN) and hyperconnectivity in salience (SN) and reward circuits, alongside increased hypothalamic– limbic coupling and reduced prefrontal control connectivity^2^. Stronger activity in the precuneus (DMN) suggests heightened self-referential focus, whereas weaker anterior cingulate engagement reflects impaired behavioral monitoring^2^. These alterations, including enhanced putamen connectivity, point to maladaptive integration of interoceptive, cognitive, and reward-related processes that may underlie habitual overeating^5,6^.

The extent to which these alterations reflect obesity-specific neural organization rather than metabolic dysfunction and eating patterns remains unclear. Emerging evidence links intrinsic network connectivity to insulin resistance^7^, metabolic syndrome^8,9^, and type 2 diabetes^10^ —even in non-obese populations—suggesting that disrupted brain connectivity may precede or exacerbate metabolic deterioration, and could be a marker signalling the impact of metabolic disruption. Importantly, brain networks which are altered in obesity, are also altered in response to eating behaviors independently of obesity profile^11^, and the genetic variants associated with obesity are impacted by eating behaviors such as emotional and disinhibited eating ^12^.

We investigated associations between large-scale functional brain organization, obesity, and metabolic health using resting-state fMRI and metabolic profiling from a large community cohort – The Nathan Kline Institute for Psychiatric Research - Rockland Sample data. We further tested how neural network profiles shape the relationship between eating behaviors and metabolic health, while controlling for lifestyle factors such as sleep quality and physical activity, and examined how network strength predicts metabolic outcomes over time. Our analyses identified a functional network that mediates the link between eating behaviour and metabolic outcomes and predicts longitudinal changes in metabolic indicators, revealing neural pathways underlying metabolic vulnerability and resilience.

## Results

### Sample characteristic and health behavioral patterns

Table 1 presents descriptive characteristics for the whole sample and mean ± SD values for each group: healthy normal weight (MHNW), metabolically healthy obese (MHO), and metabolically unhealthy obese (MUO); metabolically unhealthy normal weight (MUNW).

**Table 1.** Demographic, metabolic, and behavioural characteristics across metabolic–weight groups. Mean ± SD values are presented for each group and for the total sample (*N* = 564).

|  | **MHNW**<br>*N* = 362 | **MHO**<br>*N* = 64 | **MUO**<br>*N* = 107 | **MUNW**<br>*N* = 29 | **Total**<br>*N* = 564 |
| --- | --- | --- | --- | --- | --- |
| Age (years) | 42.04<br>(21-64) | 42.24<br>(21-64) | 46.50<br>(21-64) | 48.00<br>(25 - 63) | 42.91<br>(21-64) |
| Sex (% female) | 68 | 54 | 68 | 66 | 66 |
| Years of Education | 15.72<br>(9-23) | 15.31<br>(12-23) | 15.08<br>(12-21) | 14.97<br>(11-21) | 15.52<br>9 - 23 |
| BMI (kg/m<sup>2</sup>) | 24.48<br>(17.33 - 29.80) | 33.53<br>(30.00 - 42.99) | 33.93<br>(30.10 - 49.96) | 27.25<br>(19.49-29.61) | 27.63<br>(17.33 – 49.96) |
| Waist circumference (cm) | 82.70<br>(60.96 - 110.5) | 102.79<br>(81.30 - 147) | 111.03<br>(91.0 - 138) | 94.93<br>(77.0 - 123) | 90.35<br>60.96 – 147) |
| Glucose | 87.06<br>(47 - 118) | 90.20<br>(55-120) | 124.34<br>(68-552) | 108.45<br>(79-302) | 92.90<br>(47 -552) |
| Cholesterol (mmol/L) | 5.64<br>(2.87 - 10.96) | 5.80<br>(3.74-8.93) | 5.77<br>(3.80-12.35) | 6.0<br>(4.03-9.89) | 5.71<br>(2.87 – 12.35) |
| Triglyceride (mmol/L) | 0.98<br>(0.34-4.0) | 1.08<br>(0.38- 2.50) | 2.28<br>(0.69 - 16.68) | 2.35<br>(0.66 - 8. 44) | 1.22<br>(0.34- 16.68) |
| HDL (mmol/L) | 1.71<br>(0.77- 3.26) | 1.52<br>(0.78 - 3.0) | 1.24<br>(0.73-2.12) | 1.21<br>(0.67 - 2.10) | 1.60<br>(0.67 -3.26) |
| Systolic blood pressure (mmHg) | 113.95<br>(78 - 175) | 120.26<br>(90-166) | 133.28<br>(102-178) | 133.52<br>(98 - 178) | 118.34<br>(78 -178) |
| Diastolic blood pressure (mmHg) | 71.28<br>(41 - 101) | 78.38<br>(58-99) | 84.56<br>(62-122) | 83.76<br>(64 - 127) | 74.79<br>(41 -127) |
| **Health Behaviours** |  |  |  |  |  |
| *Eating behaviour patterns* |  |  |  |  |  |
| Disinhibition | 4.34 (2.97) | 6.77 (3.96) | 7.41 (4.07) | 5.21(3.37) | 5.21 (3.55) |
| Hunger | 3.81(2.93) | 6.33 (3.63) | 6.08(4.25) | 4.10 (3.20) | 4.38 (3.36) |
| Restraint | 8.61(5.09) | 9.38 (4.66) | 9.13 (4.51) | 8.66 (5.03) | 8.95 (4.93) |
| Sleep Quality (PSQI) | 4.74 (3.49) | 5.19 (2.84) | 6.41 (3.99) | 6.03 (3.91) | 5.81 (3.50) |
| Physical Activity (Met - Min/week Scores) | 641.87 (812.17) | 717.36 (994.63) | 654.99 (974.08) | 594.81(607.96) | 655.36 (858.87) |

*Note.* Values in parenthesis represent min – max; mmol/L = milimoles per litre; Met = Metabolic Equivalent).

The total sample (*N* = 564) had a mean age of 42.9 years (range = 21–64) and comprised 66% women. Most participants identified as White (78 %), followed by Black or African American (13 %) and Other (American Indian, Asian, or Pacific Islander; 9 %). Participants had an average BMI of 27.6 kg/m^2^ and mean waist circumference of 90.4 cm. Mean fasting glucose and cholesterol levels were 92.9 mg/dL and 5.71 mmol/L, respectively, with 25% meeting criteria for metabolic syndrome (> 2 indicators). Average systolic and diastolic blood pressure values were 118.3 mmHg and 74.8 mmHg, respectively. Regarding behavioral measures, mean disinhibition, hunger, and restraint scores were 5.21 ± 3.55, 4.38 ± 3.36, and 8.95 ± 4.93, respectively, while mean sleep quality (PSQI) score was 5.81 ± 3.50, indicating moderate sleep disturbance. Total physical activity, assessed by IPAQ, averaged 655.4 ± 858.9 MET·min/week, corresponding to approximately 2–3 hours of moderate-intensity activity weekly—consistent with current WHO recommendations.

Two-way ANOVAs were conducted to examine the effects of BMI (obesity status; normal weight vs. obese) and metabolic profile (healthy vs unhealthy) on eating behavior subscales (disinhibition, hunger and restrain), physical activity and sleep quality index (*N* = 551). Obesity status significantly influenced disinhibition (*F*(1,542) = 31.67, *p* < 0.001, *η*^*2*^ = 0.055) and hunger (*F*(1,542) = 17.73, *p* < 0.001, *η*^*2*^ = 0.032), with higher scores among individuals with obesity. In contrast, poor metabolic health was associated only with poorer sleep quality (*F*(1,547) = 8.34, *p* = 0.004, *η*^*2*^ = 0.015). No significant main or interaction effects were observed for eating restraint or physical activity measures (all *p* > 0.11).

### Brain network signatures of metabolic health and obesity

Group differences in rsFC were based on group obesity x metabolic health comparison. No significant main effect of metabolic health or obesity-related differences in rsFC was observed.

We conducted pairwise comparisons to examine the brain correlates associated with different metabolic profiles in individuals with obesity. The analysis focused on subgroups differences using one tailed t-test, while correcting for the number of significant nodes of a network (i.e. family-wise correction at the network level). Among the four subgroups, two pairwise comparisons showed significant group differences in rsFC: MHO vs MUO, revealing network signatures of metabolic health and MHNW vs MHO, revealing network signatures of obesity.

Comparing rsFC between MHO and MUO revealed significantly higher between-network connectivity in the MUO group, originating from the somatomotor (SMN) network to the default mode network (DMN) and frontoparietal control network (CN), with reciprocal connections among the parietal and prefrontal regions in DMN and CN (Fig. 1A &C, Supplementary Table S1). A few nodes in the visual pathway (visual network (VN), dorsal attention network (DAN) and salience/ventral attention network (SVAN)) were also involved. By contrast, the MHO group showed significantly higher within-network connectivity within the SMN (both unilateral and bilateral connections), plus DAN – limbic network (LN) coupling (Fig. 1A, Supplementary Table S2). Of note, both SMN and DAN showed significantly enhanced connections with the right amygdala.

**Figure 1.**
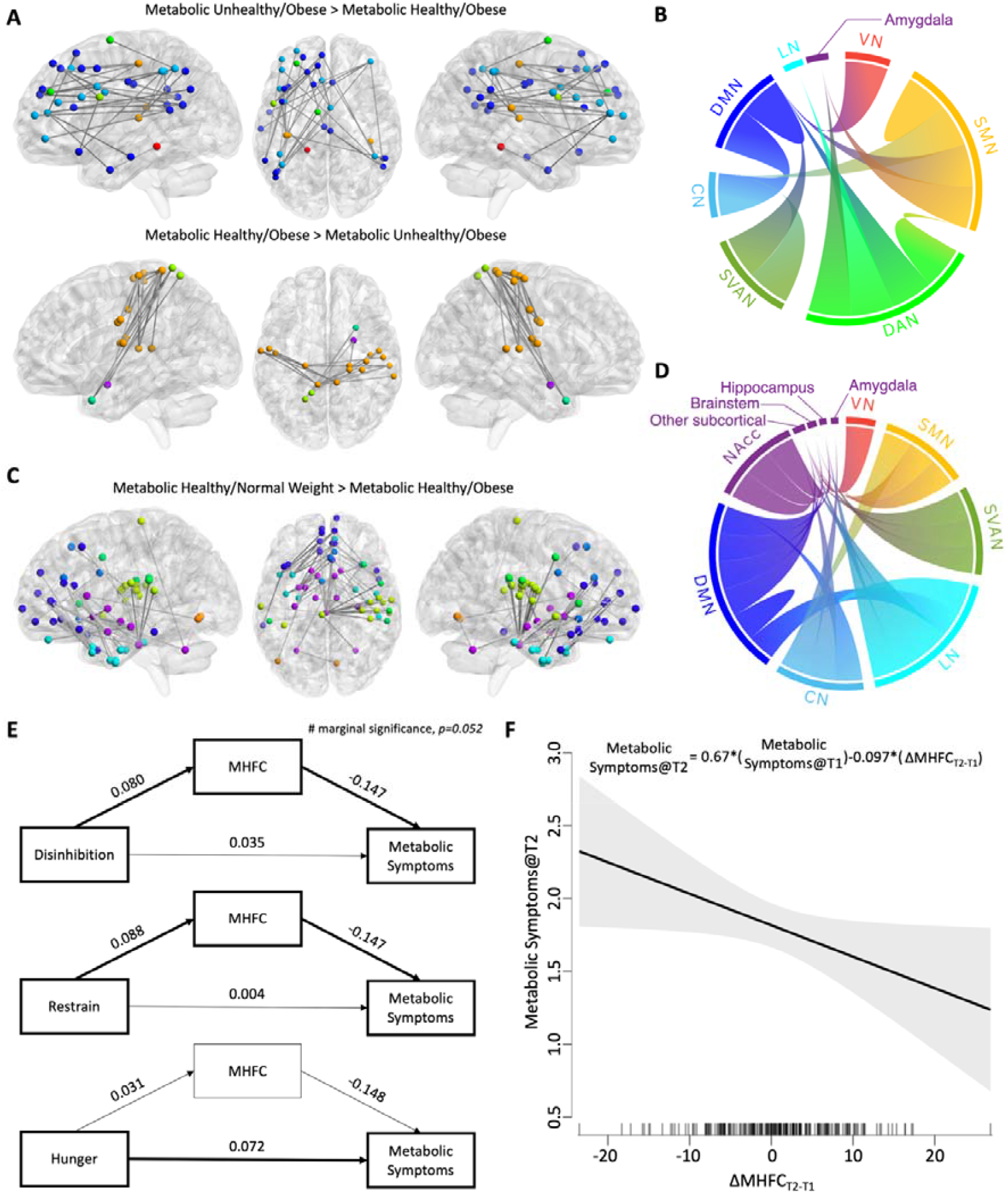
Functional connectome (using the Schaefer 400 parcels + 19 subcortical regions atlas) comparisons between **A)** the Metabolic Healthy/Obese (MHO) and the Metabolic Unhealthy/Obese groups (MUO); **C)** the Metabolic Healthy/Normal Weight (MHNW) and the Metabolic Healthy/Obese (MHO) groups, p<0.05 FWE corrected at the network level**; B), D)** Chord diagram depicting the respective edges presented in **panel A, C**, group by the Yeo et al. (2011) 7 network scheme (VN: Visual network; SMN: Somatosensory Motor Network; DAN: Dorsal Attention Network; SVAN: Salience/Ventral Attention Network; LN: Limbic Network; CN: Frontoparietal Control Network; DMN: Default Mode Network; Purple nodes: subcortical regions); **E)** Path models demonstrating the significant role of MHFC (all edges presented in **panel A**) mediating restrained eating and metabolic symptoms, as well as marginally non-significant mediation between disinhibited eating and metabolic symptoms; **F)** using available longitudinal data from NKI, the change in MHFC network strength significantly predicts metabolic symptoms at T2, after controlling for the number of metabolic symptoms at T1 and other confounding factors (age, sex, education, BMI). An increase in MHFC network strength is associated with reduced metabolic symptoms. Regression equation is provided on top of the plot.

The observed differences between MUO and MHO reflect distinct rsFC patterns linked to metabolic health status. Degraded metabolic health is associated with increased connectivity among CN and DMN. SMN, likely representing a loss of functional segregation in the MUO group manifest as excessive influence of the sensorimotor system on self-referential and interoceptive processes. The MHO group showed stronger within network connectivity among the SMN and effective functional integration of DAN-LN for the processing of emotional and interoceptive cues.

Comparing rsFC between MHNW and MHO groups revealed increased connectivity between SMN, cerebellum and SVAN with brainstem as the central hub, as well as connections between LN, DMN, CN and reward related subcortical brain regions (bilateral Nucleus Accumbens, Hippocampi and left amygdala) (Fig. 1B & D, Supplementary Table 3). The differences between MHNW and MHO reflect distinct rsFC patterns linked to obesity status. The rsFC pattern suggests that higher-level cortical networks (DMN, SVAN, CN) are stronger coupled with subcortical hubs that underlie emotion, reward, and arousal processing potentially associated with calorie intake and obesity. Robustness of the metabolic health-related and obesity-related rsFC differences are examined and presented in Extended Data (Fig. S1 & S2).

### Network Configuration as mediator of eating behavior and metabolic health

Our findings reveal two distinct patterns of functional brain organization: one identified as metabolic health-related functional connectivity (MHFC; MHO vs. MUO) and another identified as obesity-related functional connectivity (OBFC; MHNW vs. MHO). We sought to determine whether these two network configurations contribute to the association between eating behaviors and metabolic health. To address this, we computed MHFC and OBFC network strengths for each participant (n=500, disregarding their grouping in previous analysis) and tested their mediating roles in the relationship between eating behaviors and metabolic health indicators. The number of metabolic health indicators was quantified according to the National Cholesterol Education Program Adult Treatment Panel III (NCEP ATP III) criteria for metabolic syndrome ^13^.

For each sub-domain of eating behavior patterns (i.e., disinhibited, hunger, and restrain), we tested their respective direct effect in predicting the number of metabolic symptoms, as well as their indirect effect mediated by one of the rsFC configurations (MHFC or OBFC). In all mediation models age, sex, level of education and BMI are entered as covariates.

Our findings revealed that MHFC network strength significantly mediated the relationship between restrained eating and metabolic symptoms (Fig. 1E). We replicate this effect with data from the same cohort measured at a second timepoint (1^st^ follow up, n=160, Extended Data Fig. S3). A marginally non-significant mediating effect for the association between disinhibition and metabolic symptoms. However, this trend could not be observed using the data from the second timepoint. OBFC network strength showed no significant mediating effect between any domains of eating behavior and metabolic health, but on models with BMI as the outcome (see Extended Data Fig. S4-S5).

Our mediation analyses suggests that networks of cognitive control and attention to emotional and interoceptive cues that characterize the MHFC, could be subserving adaptive eating by executive control functions involving emotional states, food cues, and interoceptive satiety signals, playing a significant role in the relationship between eating restrain, i.e. conscious control of food intake to control body weight and metabolic health. This line of thought fits equally well to disinhibition, which shows the opposite pattern (i.e., disinhibition as the eating pattern characterized by loss of executive control over food consumption) to restrained eating, albeit with less robust results. Of note, the mediation models are tested on the whole sample disregarding their subgroups, meaning the observed effect is not restricted in the obese subgroups.

### Effects on network configurations on metabolic health over time

Although the mediation analyses seem to suggest causal relationship, the models are cross-sectional in nature, and the observed associations are by no means causal. Therefore, we conducted a longitudinal analysis using the NKI’s available follow-up data (n=228). Using hierarchical regression analysis, we tested whether differences in individual’s network strength in MHFC and OBFC predicted changes in metabolic indicators over time, while controlling for baseline levels of metabolic health, age, sex, level of education, and concurrent BMI measured at follow-up.

Hierarchical regression showed that baseline covariates explained 57.5% of the variance in metabolic symptoms (*F*(16, 214) = 18.06, *p* <.001). The inclusion of changes in metabolic network strength (ΔMHFC) modestly improved model fit (ΔR^2^ =.009, *F*(1, 213) = 4.47, *p* =.036), with reduced network strength predicting higher metabolic indicators at follow-up (β = −.097, Fig. 1F). This reflects that a positive change in MHFC but not OBFC network strength predicts the reduction of metabolic indicators, and this effect withstood the inclusion of metabolic symptoms at baseline and covariates.

## Results summary

This study identifies two distinct resting-state network configurations, one delineating metabolic health (MHNC) and another obesity (OBNC). The MHFC configuration—linking regions of the default mode, salience, and frontoparietal control networks— mediated the relationship between eating behavior and metabolic indicators independently of BMI, a finding that was replicated in data acquired at a separate time point.

Longitudinal analyses demonstrated that stronger MHFC at baseline predicted improved metabolic profiles at follow-up, underscoring its potential role as a marker of metabolic and brain function resilience.

## Discussion

Our findings identify distinct patterns of resting-state functional connectivity that differentiate obesity- and metabolic health–related brain organization in a community dwelling population of young and middle-aged adults. To our knowledge, this is the first study to systematically characterize the brain network organization associated with both obesity and metabolic function. These results also support and extend previous neuroimaging work showing that obesity is characterized by altered large-scale networks governing reward, self-referential processing, and cognitive control ^3,5,14^. We provide evidence that disruptions in brain network organization associated with the presence of metabolic syndrome indicators (i.e., defined by established clinical criteria) occurs independently of obesity status.

Our study provides novel insights into the distinct functional brain organization associated with both metabolic health and obesity. Recognizing this distinction is essential, as disruptions in glucose and lipid metabolism and obesity are mechanistically associated with changes in brain structure and function ^8,10,15–17^. Although a growing body of evidence suggests that individuals with obesity can exhibit distinct metabolic profiles —ranging from metabolically healthy to metabolically unhealthy phenotypes ^18,19^ —previous neuroimaging studies have typically focused on either obesity or metabolic dysfunctions such as metabolic syndrome or diabetes, without directly examining both factors within the same framework. Previous studies addressing the effects of metabolic dysfunction on resting-state functional connectivity have shown that metabolic syndrome and related indicators—such as obesity, dyslipidemia, and hyperglycemia—are associated with widespread reductions in network coherence, particularly within the default mode, executive control, and salience networks ^8,20^. In parallel, studies examining cardiovascular health and insulin resistance have demonstrated that higher body mass index and reduced insulin sensitivity are linked to altered connectivity within the default mode, orbitofrontal, and insular networks, further implicating disrupted metabolic and vascular signalling ^4,17^.

Our results highlight a shift from efficient segregation toward excessive crosstalk between the somatomotor (SMN), default mode (DMN), and frontoparietal control networks (CN) in metabolically unhealthy obesity (MUO). This reorganization may reflect reduced top-down modulation of interoceptive and emotional signals, resulting in heightened sensitivity to bodily and food-related cues. In contrast, metabolically healthy obesity (MHO) was characterized by stronger intra-network coherence within the somatomotor system and enhanced coupling between attention and limbic networks, suggesting preserved integration of interoceptive and emotional processing. This pattern partially resembles those from previous rsFC studies on individuals with metabolic syndrome ^9,20^, with abdominal obesity ^21^, and with type 2 diabetes patients ^17^.

Eating behaviors are closely linked to obesity and metabolic outcomes ^22,23^. In line with recent evidence demonstrating that individual differences in eating behavior traits are reflected in resting-state connectivity profiles independent of body weight ^11^, we found that the metabolic health functional connectivity (MHFC) network mediates the association between eating restraint and metabolic health. This network, encompassing regions involved in executive control and interoceptive regulation, suggests that successful control over eating and craving operates through neural mechanisms integrating self-regulation with bodily state monitoring. Notably, the mediation effect of the MHFC network was observed across obesity status, indicating a broad, non-specific role of this circuitry that extends beyond differences in body weight or metabolic syndrome.

Previous studies have linked resting-state functional networks to eating behaviours in individuals with and without obesity but did not examine metabolic health profiles. Altered connectivity within executive control networks in individuals with abdominal obesity has been linked to reduced self-regulation of eating, suggesting that visceral fat accumulation may impair neural systems supporting cognitive control over food intake ^21^. Similarly, heightened caudate–hippocampal connectivity correlates with emotional eating, implicating reward– memory interactions in hedonic food intake ^24^. Altered interactions among resting-state networks**—**particularly involving the salience, executive control, and default mode systems— have also been reported in individuals with obesity, pointing to disrupted coordination between control and reward processes that may underlie dysregulated eating behaviour ^25^. Together, these findings highlight functional network alterations associated with eating behaviour, while our mediation analysis extends this framework by linking network configuration to both eating restraint and metabolic health. Our results suggest a shared neural pathway integrating behavioural control and metabolic regulation.

Our longitudinal analyses further examined whether metabolic health network configuration predicts metabolic outcomes over time. This analysis revealed that decreases in MHFC strength at baseline were associated with worsening metabolic indicators, suggesting a bidirectional relationship between functional network integrity and metabolic regulation. This is the first study that identifies brain network configurations at baseline to predict changes in metabolic indicators. These findings echo previous cross-sectional reports linking insulin resistance and metabolic syndrome to disrupted prefrontal and limbic network coupling, particularly within default mode, salience, and executive control systems ^9,14,20^.

A few studies have examined whether baseline functional network configuration predicts subsequent weight loss, hormonal changes, or eating behaviour. For example, in healthy women, connectivity between the ventral striatum and orbitofrontal cortex predicted future weight gain and increased leptin levels, implicating reward-system hyperconnectivity in metabolic vulnerability ^26^. Among individuals with obesity undergoing bariatric surgery, stronger preoperative coupling within the default mode and executive control networks predicted greater postoperative weight loss ^27^. In a behavioural intervention for older adults, baseline connectivity in prefrontal and salience regions predicted weight-loss success ^28^. Although these studies did not comprehensively assess longitudinal changes in metabolic markers, their findings converge to suggest that the integrity of reward, control, and interoceptive networks at baseline shapes subsequent behavioural adaptation and weight loss. Together, they indicate that alterations in large-scale connectivity not only accompany but may also precede metabolic decline, behavioral and weight change, highlighting a potential neural pathway through which dysregulated brain signalling impacts metabolic health.

Several limitations warrant consideration. While the inclusion of longitudinal data enhances causal interpretation, the current analyses cannot definitively establish the directionality of brain–metabolic relationships or eliminate the possibility of residual confounding. A range of physiological, behavioral, and environmental variables not captured in this study—such as dietary composition, hormonal variability, medication use, and chronic stress—may have influenced both metabolic outcomes and brain network organization. Furthermore, reliance on self-reported measures of eating behavior, sleep quality, and physical activity introduces potential reporting bias. Future research integrating detailed metabolic, hormonal, and lifestyle profiling with multimodal neuroimaging will be essential to confirm these results and clarify the neural pathways linking functional brain organization with metabolic health.

In summary, this study identifies a reproducible brain connectivity signature associated with metabolic health that predicts longitudinal changes in metabolic indicators. The metabolic health network mediated the relationship between eating behavior patterns and metabolic outcomes, suggesting that individual differences in functional connectivity supporting cognitive control and interoceptive regulation contribute to metabolic health. This effect was replicated at a follow-up time point, highlighting its robustness. Finally, longitudinal analyses showed that baseline network strength predicted changes in metabolic indicators over time, underscoring the important role of functional brain organization as potential marker of resilience or vulnerability in the progression of metabolic dysfunction.

## Methods

### Participants

The Nathan Kline Rockland Sample includes participants from Rockland County^29^, a population of 311,687 located 24 km northwest of New York City. The racial demographics of Rockland County are 72.5% White, 17.4% African American, 6% Asian, 1.2% Native American, 0.34% Native Hawaiian, 2.5% Other Race. This resembles those of the United States Census (U.S. Census Bureau, 2009). Participants recruitment was mainly through flyers posted at schools, shopping malls, community centers, and other locations within Rockland County. We examined data from a subset of the adult participants (*N*= 564), ages 21 to 64 years (*M* = 42.86, SD = 13.46), who had complete metabolic data and underwent semi-structured diagnostic psychiatric interviews, including eating behavior, physical activity, and sleep quality questionnaires, administered by the NKI research team. The subset included 189 males and 375 females (see Table 1).

The present study included individuals with complete medical history, metabolic and anthropometric data. Participants were excluded if they reported: (1) presence of neurological or cerebrovascular conditions; (2) untreated severe depressive symptoms, and substance abuse; (3) history of cardiovascular disease or brain injury (e.g., stroke), epilepsy, seizures; (4) psychotropic medication use in the past three months; and (5) bipolar disorder, schizophrenia, or psychotic disorder; (6) history of ischemic attack. Individuals with missing structural MRI or resting state fMRI data, significant motion artefact, or moderate to severe blurring artefact were further excluded.

All methods were carried out in accordance with the Institutional Review Board (IRB) at the Nathan Kline Institute. Approval was obtained for this project at the Nathan Kline Institute IRB (Phase I #226781 and Phase II #239708) and at Montclair State University (Phase I #000983A and Phase II #000983B). Written informed consent was obtained for all study participants.

## Measurements

### Measurement of laboratory and anthropometric parameters

All data on laboratory and anthropometric parameters, as well as medical history were downloaded from the NKI database.

### BMI group

Participants were categorized into BMI groups: normal weight (18.5-24.9), overweight (25.0-29.9), and obesity (>= 30.0). Anthropometric and blood pressure (BP) measurements were conducted comparably in both cohorts. Blood samples were analysed for triglyceride, total cholesterol, high-density lipoprotein cholesterol, and glucose.12,13. Race/ethnicity and prevalent diabetes were self-reported; current medication was self-reported and cross-checked on medical reports.

### Metabolic health

A cluster of metabolic and cardiovascular risk factors has been used in to define individual’s metabolic health. Metabolic health was defined according to the National Cholesterol Education Program Adult Treatment Panel III (NCEP ATP III) criteria. Individuals meeting fewer than three of the five criteria (elevated waist circumference, triglycerides, blood pressure, fasting glucose, and reduced HDL cholesterol) were classified as metabolically healthy, whereas those meeting three or more criteria were considered metabolically unhealthy. Individuals are considered healthy if 2 or fewer criteria are present: 1) waist circumference, 102 cm (men) or greater than 88 cm (women); 2) BP greater than or equal to 130/85 mm Hg or 3) using BP-lowering medication; 4) triglyceride level greater than or equal to 150 mg/dL or 5) using lipid-lowering medication; 6) high-density lipoprotein cholesterol (HDL) level less than 40 mg/dL (men) or less than 50 mg/dL (women); and 7) fasting glucose level greater than or equal to 110 mg/dL or 8) prevalent diabetes ^23^. To quantify individual subject’s metabolic health, the total number of metabolic symptoms was used.

Metabolically healthy and unhealthy individuals were further grouped according to their obesity status (obese = BMI >= 30), resulting in a total of four subgroups: metabolically healthy normal weight (MHNW), metabolically healthy obese (MHO), metabolically unhealthy normal weight (MUNW), and metabolically unhealthy obese (MUO) subgroups. We adjusted all the analysis for the following known confounders (self-reported with predefined categories to choose from): age, sex (male, female), educational level (in years), and BMI

## Behavioral measures

### Eating behavior patterns

We used the Three Factor Eating Questionnaire (TFEQ)^30^, a validated 51-item instrument designed to measure three dimensions of eating behavior: disinhibition, which reflects the tendency to lose control over eating in the presence of palatable foods or emotional stress; hunger which reflects subjective feelings of hunger and food cravings; and restraint which reflects conscious control or restriction of food intake to control body weight. Examples of scale items include for disinhibition: “Sometimes when I start eating, I just can’t seem to stop”, for hunger: “I often feel so hungry that I have to eat something right away” and for restraint: “I often stop eating when I am not really full as a conscious means of limiting the amount that I eat”. The questionnaire provides 20 items for restraint, 16 for disinhibition, and 15 for hunger score. The instrument contains 36 yes/no items and 15 on a 1-4 response scale. Each eating behavior dimension is calculated by summing responses to items for each category. Higher scores denote higher levels of disinhibition, hunger, and restraint. The TFEQ demonstrates adequate reliability and validity for assessing eating behaviors in young and older adults^31^. Previous studies reporting on the TFEQ using the NKI dataset have reported similar population scores^32–34^, and have shown adequate reliability and internal consistency for the NKI sample^35^. Adequate internal consistency was also obtained for the present sample: disinhibition (α = 0.76), hunger (α = 0.71), and restrain (α = 0.74).

### Sleep quality

The Pittsburgh Sleep Quality Index (PSQI)^36^ consists of a self-report questionnaire that assesses sleep quality and disturbances over the preceding month. It consists of 19 items, which are combined into seven component scores which are summed to obtain a total sleep quality score ranging from 0 to 21, where higher scores reflect poorer sleep quality. A global PSQI score >5 is conventionally used to differentiate “poor” sleepers from “good” sleepers. The PSQI has demonstrated good reliability and validity in both clinical and research populations.

### Physical activity

The International Physical Activity Questionnaire (IPAQ)^37^ is a validated self-report instrument designed to quantify physical activity during the previous 7 days. It assesses the frequency (days per week) and duration (minutes per day) of vigorous, moderate, and walking activities. In line with previous studies, we calculated a combined total MET score by summing the MET-minutes per week from vigorous, moderate, and walking activity. This score reflects the total volume of physical activity performed per week, expressed in MET-minutes/week, with higher scores indicating greater activity.

## MRI analysis

### MRI data acquisition and processing

High-resolution T1-weighted structural images and resting state functional images were downloaded for functional brain network analyses. High-resolution T1-weighted anatomical images were acquired for co-registration with the following sequence parameters: field of view (FOV) = 250 × 250 mm, voxel size = 1 mm3, repetition time (TR) = 1900 ms, echo time (TE) = 2.52 ms, TI = 900 ms, flip angle = 9°, matrix = 256 × 256, phase encoding A >> P, GRAPPA accel. factor = 2, and sequence duration = 4:18 min. Resting state BOLD data were acquired using the following sequence parameters: gradient echo planar images, FOV = 222 × 222 mm, voxel size = 3.00 mm3, TR/TE = 645/30 ms, number of slices = 40, flip angle = 60°, matrix = 74 × 74, phase encoding A > > P, Multiband factor = 5, and sequence duration= 9:37 min^38^. The one-day protocol included both neuropsychological testing and MRI.

### MRI Preprocessing and Functional connectivity analysis

Resting state BOLD data were preprocessed with SPM12 (http://fil.ion.ucl.ac.uk/spm/) using standard preprocessing pipelines, which includes motion correction, coregistration with T1 structural scan, unified segmentation of T1 structural scan to obtain normalization parameters to the MNI space, normalizing the BOLD data, and spatial smoothing with 6mm FWHM kernel. The resultant images were further preprocessed with DPABI toolbox^39^ for bandpass filtering between 0.01-0.08Hz, linear trend removal, as well as regressing out motion parameters for resting state functional connectome analyses.

We used the Schaefer 400 parcels brain atlas^40^ in combination with the Freesurfer subcortical region-of-interests to parcellate whole brain into 419 regions. Within the voxels from each brain region we calculated the averaged timecourse and use it computed pair-wise correlations (Fisher’s z) between all 419 regions to create a 419 x 419 connectivity matrix for each subject. Statistical comparisons between groups were performed with the Network Based Statistics (NBS) toolbox^41^. To control for network level false positives, statistical significance was determined by permutation test (5000 permutations) to obtain the null distribution of randomly connecting network, thresholded at p=0.0001 for individual edges. This procedure is equivalent to calculating the Family-wise Error (FWE) correction at the network level, *p*_*FWE*_*>0*.*05*.

## Data Availability

https://rocklandsample.org/

