## Extended Data for "Metabolic health specific functional connectivity signatures in the human brain"

Additional analyses

*rsFC robustness check:* To check for the robustness of our brain network signatures of metabolic health and obesity, we performed additional analyses by 1) restricting our sample to subjects aged 40 or above, and 2) restricting our sample to subjects with data on their serum level of C-Reactive Protein (CRP), a biomarker of low grade inflammation and metabolic dysfunction, as an independent validation of the grouping.

We reasoned that by restricting our sample to subjects aged 40 or above (n=302), we should observe more robust effects as more metabolic symptoms is expected after middle age. We replicated the brain network differences between MHO and MUO in our restricted sample, characterized by the increased within-network connectivity of the SMN and DAN in the MHO group. The same pattern of increased within-network connectivity is also observed in the MHO group when comparing to the MHNW group, and appeared as a main effect of metabolic health. We also observed a marginally non-significant difference pattern of network connectivity for the MHNW > MHO contrast, which is similar to but less robust than our main analysis. The key connection observed in the main analysis, namely coupling between NAcc, Hippocampus, brainstem & DMN are replicated.

We further employ serum level of CRP as an independent biomarker for metabolic dysfunction to validate our findings. Since not all NKI subjects we used in the main analysis have CRP measured, this selection reduced the total sample size to 178 subjects (MHNW n=119, MHO n=32, MUNW n=10, MUO n=17). 2 x 2 ANOVA examining metabolic health x obesity on CRP revealed a significant main effect of obesity (F(1,174) = 8.593, p=0.004) and a significant metabolic health x obesity interaction (F(1,174) = 3.849, p=0.05), providing external validity of our grouping criteria. Post hoc test revealed significant differences CRP_MUNW_ > CRP_MHNW_, while both CRP_MHO_ & CRP_MUO_ are elevated. With this reduced sample we performed the same group comparisons on rsFC, and we replicated the previously observed pattern of increased within-network connectivity in SMN and DAN (Fig. S2A). Similar connectivity pattern was also observed in the MHNW > MUNW contrast, although it’s less robust and only marginally non-significant. We also observed marginally non-significant pattern of increased coupling between NAcc, Hippocampus and DMN in the MHNW > MHNW contrast, but this result bear no direct replication to our main findings.

The repeated observation of increased within-network connectivity of the SMN and DAN might be a key signature associated with metabolic health.

*Replicability of the mediation role of MHFC:* Utilizing NKI’s longitudinal data, we replicate the mediation analyses with data from the follow-up assessment (T2, n=160). Among the three subscales of FFQ, restrained eating is mediated by individual’s strength of MHFC to predict metabolic symptoms, all measured at T2 (Fig. S3). This replicates the main findings on restrained eating presented in the main analysis (Fig. 1E). The marginally non-significant mediation of MHFC between disinhibited eating and metabolic symptoms, as well as the direct effect of hunger on metabolic symptoms, were not replicated.

*Characterizing the OBFC:* Although the OBFC strength appears to play no significant role in mediating eating behaviours and metabolic symptoms (Fig. S4), we observed its significant role in mediating the association between eating behaviours and BMI. Specifically, OBFC network strength significantly mediates between restrained eating (Fig. S5A) and BMI, and this mediation is replicated at T2 (n=160, Fig. S5B). Similar to our main analysis on MHFC, we also tested a longitudinal regression model with BMI measured at T2 as outcome (n=228). After controlling the effects of age, sex, level of education, BMI measured at baseline, and their interactions, changes in OBFC and MHFC network strength between T1 and T2 both significantly predicts BMI measured at T2:

BMI_T2_ = -0.081*ΔOBFC_T2-T1_ -0.081*ΔMHFC_T2-T1_	(covariates omitted)

Altogether the mediation analyses and longitudinal regression analysis show that the OBFC carries biological relevance specifically targeting outcome measures related to obesity, but not metabolic health.

**Figure S1** Results of group comparisons when we restrict our analyses to subjects aged above 40 (n=302; MHNW: n=194, MHO: n=51, MUNW n=21, MUO n=36).

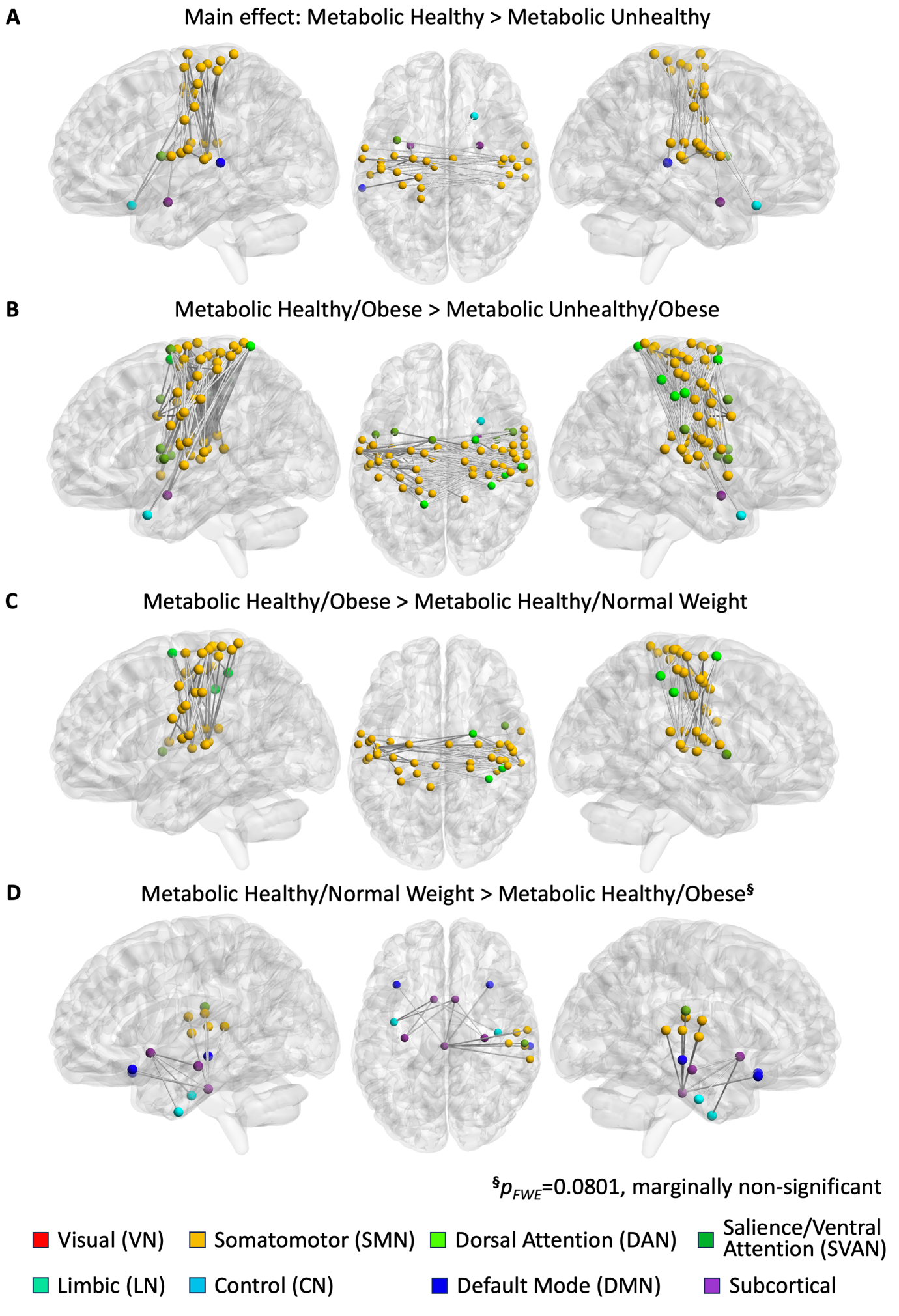

**Figure S2** Results of group comparisons when we restrict our sample to subjects with concurrent serum level of CRP as additional biomarker of low grade inflammation and metabolic dysfunction (n=178; MHNW n=119, MHO n=32, MUNW n=10, MUO n=17).

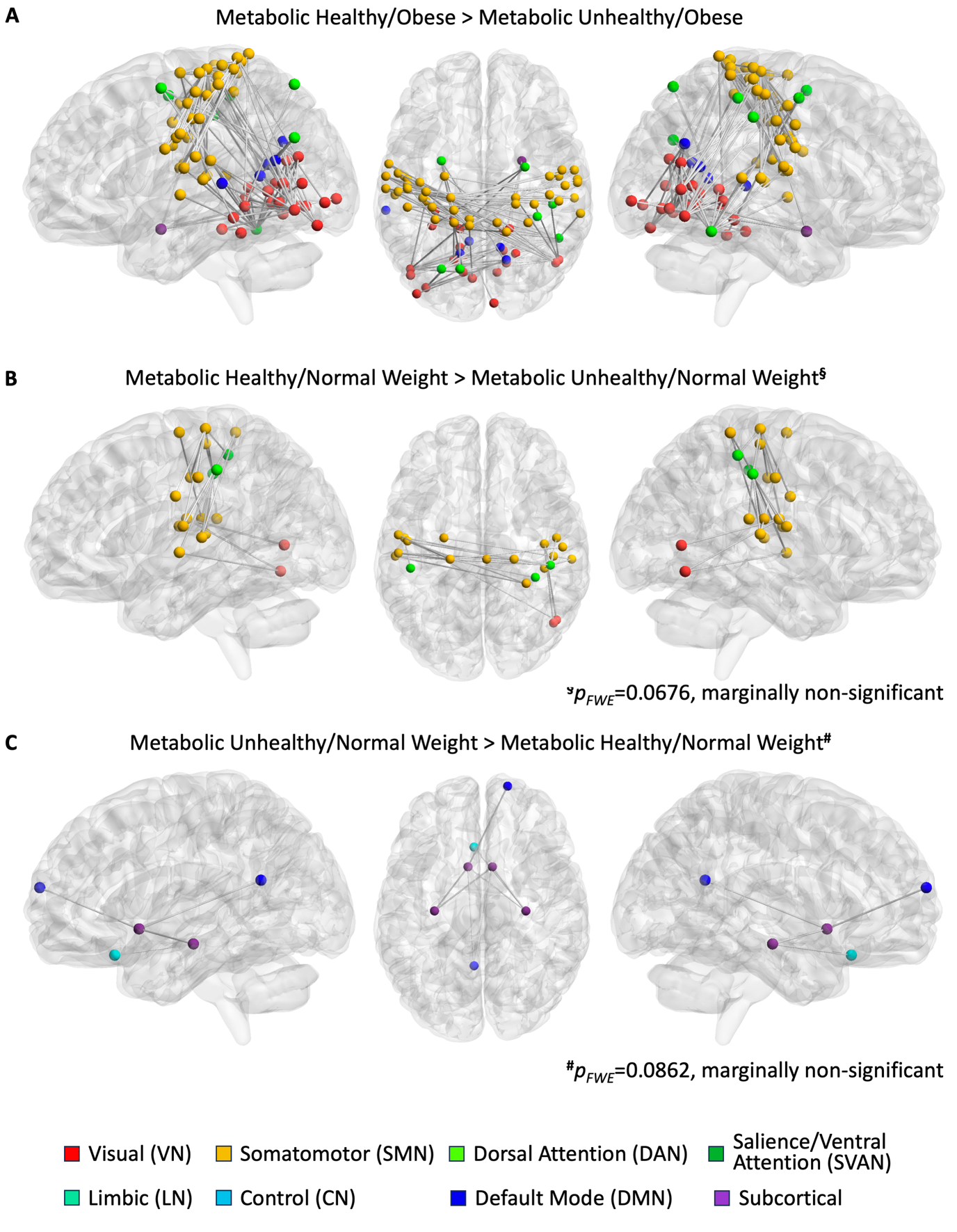

**Figure S3** Mediation effects of MHFC between three FFQ subscales and metabolic symptoms using the following timepoint of longitudinal data from NKI. Significant mediation of MHFC on restrained eating was observed, replicating our main mediation results

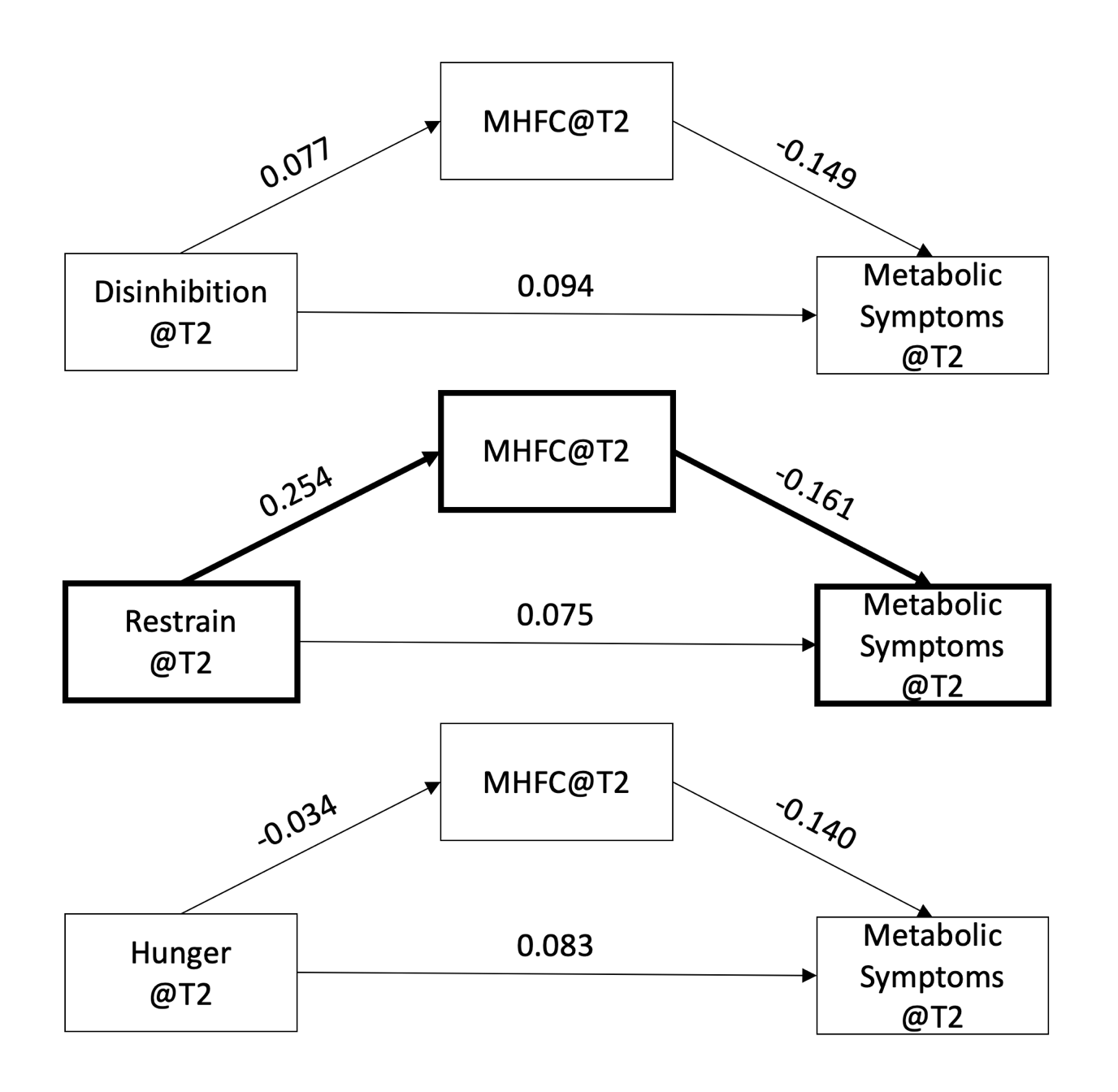

**Figure S4** OBFC showed no significant mediation between FFQ subscales and metabolic symptoms

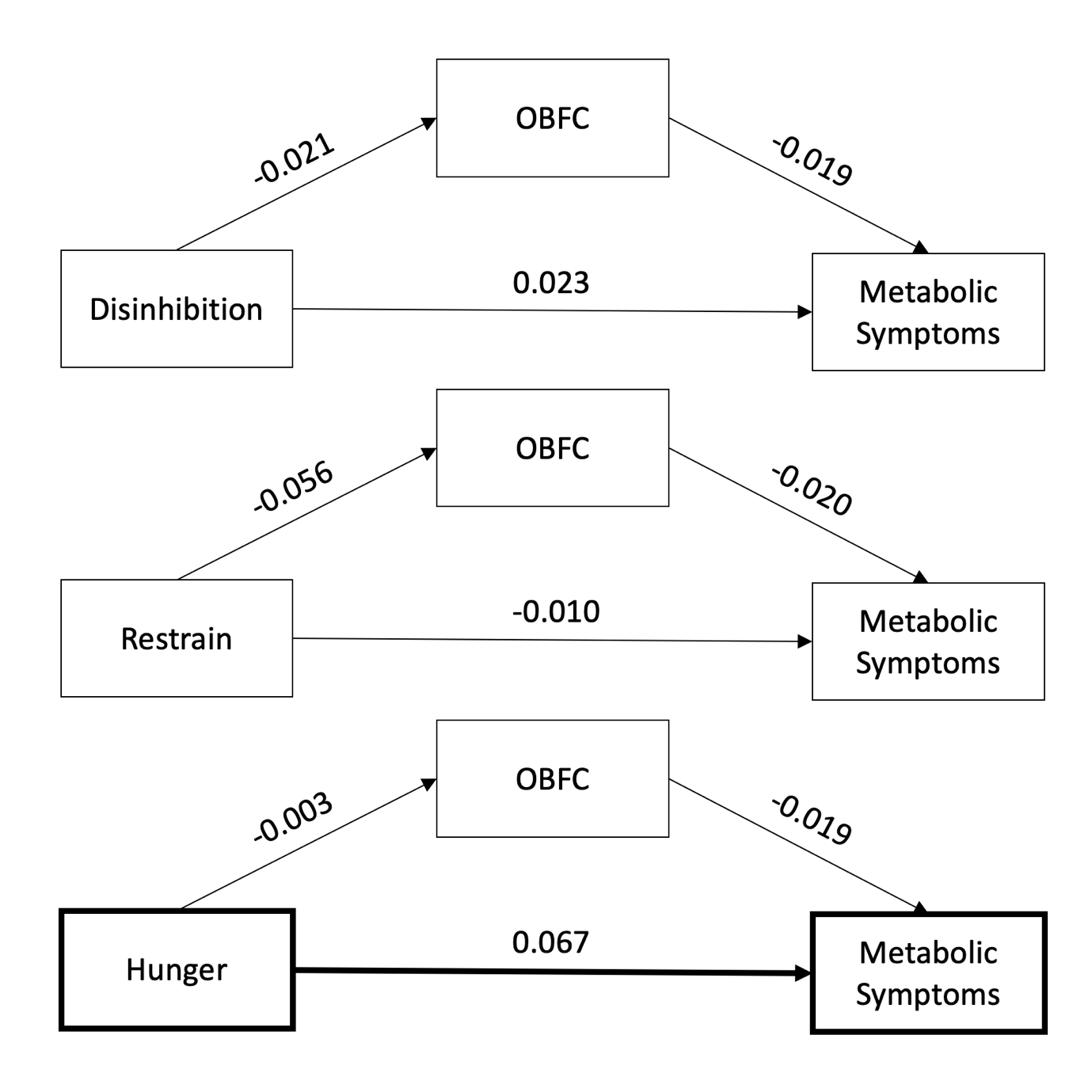

**Figure S5** OBFC showed significant mediation effect between retrained eating and BMI, which was replicated on the following timepoint of NKI longitudinal data

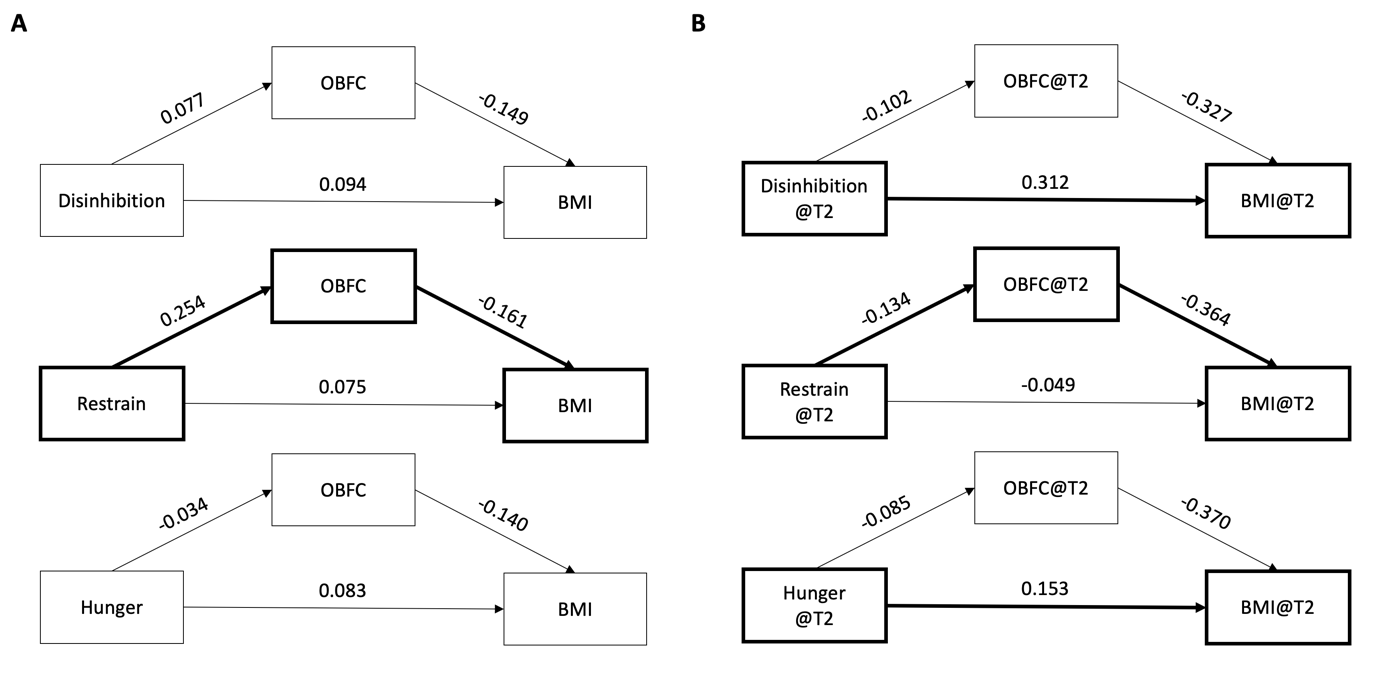
