## Supplementary material for "Metabolic health specific functional connectivity signatures in the human brain": Supplemenatry Tables

**Supplementary Information**

**Table S1**

List of connections between brain areas for the Metabolic Unhealthy (MUO)/Obese > Metabolic Healthy/Obese (MHO).

| from | to |
| --- | --- |
| LH_Vis_7 | LH_Default_Temp_1 |
| LH_SomMot_3 | LH_Cont_Par_5 |
| LH_SomMot_3 | LH_Default_Par_7 |
| LH_SomMot_3 | RH_Default_Par_4 |
| LH_SomMot_22 | LH_Cont_PFCl_6 |
| LH_SomMot_22 | LH_Cont_PFCmp_1 |
| LH_SomMot_22 | LH_Default_PFC_15 |
| LH_SomMot_22 | LH_Default_PFC_17 |
| LH_SomMot_22 | LH_Default_PFC_20 |
| LH_DorsAttn_PrCv_1 | LH_Default_PFC_1 |
| LH_DorsAttn_PrCv_1 | LH_Default_PFC_13 |
| LH_DorsAttn_PrCv_1 | LH_Default_PFC_18 |
| LH_SalVentAttn_PFCl_1 | LH_Default_PFC_19 |
| LH_SalVentAttn_Med_7 | LH_Cont_Par_5 |
| LH_Cont_Par_5 | LH_Default_pCunPCC_9 |
| LH_Cont_Par_6 | LH_Default_PFC_1 |
| LH_Cont_Par_6 | LH_Default_PFC_13 |
| LH_Cont_PFCl_1 | LH_Default_Par_1 |
| LH_Cont_PFCl_4 | LH_Default_Par_1 |
| LH_Cont_PFCl_5 | RH_Default_Par_3 |
| LH_Cont_PFCl_7 | LH_Default_Par_1 |
| LH_Cont_PFCl_7 | LH_Default_PFC_13 |
| LH_Cont_PFCl_7 | LH_Default_PFC_18 |
| LH_Cont_PFCl_7 | RH_Default_Par_3 |
| LH_Default_Temp_1 | RH_Cont_PFCl_5 |
| LH_Default_Temp_2 | RH_Cont_PFCl_5 |
| LH_Default_Par_2 | RH_Cont_Cing_2 |
| LH_Default_Par_2 | RH_Default_PFCdPFCm_6 |
| LH_Default_Par_4 | RH_Cont_PFCl_5 |
| LH_Default_PFC_13 | RH_Cont_Par_5 |
| LH_Default_PFC_18 | RH_Cont_Cing_2 |
| LH_Default_PFC_19 | LH_Default_pCunPCC_9 |
| LH_Default_PFC_19 | RH_Cont_Cing_2 |
| LH_Default_PFC_23 | RH_Cont_PFCl_5 |
| LH_Default_pCunPCC_9 | RH_Default_Par_3 |
| RH_SomMot_10 | RH_Cont_Par_5 |
| RH_Cont_Par_5 | RH_Default_Par_1 |
| RH_Cont_PFCl_5 | RH_Default_Par_3 |
| RH_Default_Par_3 | RH_Default_PFCdPFCm_6 |
| RH_Default_Par_3 | RH_Default_pCunPCC_7 |

**Table S2**

List of connections between brain areas for the Metabolic Healthy/Obese (MHO) > Metabolic unhealthy/Obese (MUO) connections.

| from | to |
| --- | --- |
| LH_SomMot_8 | LH_SomMot_32 |
| LH_SomMot_8 | RH_SomMot_32 |
| LH_SomMot_13 | LH_SomMot_31 |
| LH_SomMot_13 | LH_SomMot_32 |
| LH_SomMot_13 | RH_SomMot_32 |
| LH_SomMot_14 | LH_SomMot_31 |
| LH_SomMot_31 | RH_SomMot_7 |
| LH_SomMot_31 | RH_SomMot_19 |
| LH_SomMot_32 | RH_SomMot_7 |
| LH_SomMot_32 | RH_SomMot_8 |
| LH_SomMot_32 | RH_SomMot_11 |
| LH_SomMot_32 | RH_SomMot_26 |
| LH_SomMot_32 | RH_SomMot_29 |
| LH_SomMot_32 | RH_SomMot_34 |
| LH_DorsAttn_Post_16 | RH_Limbic_TempPole_5 |
| LH_DorsAttn_Post_17 | RH_SomMot_9 |
| LH_DorsAttn_Post_17 | RH_Limbic_TempPole_5 |
| LH_DorsAttn_Post_17 | RH_Amyg |
| RH_SomMot_7 | RH_SomMot_32 |
| RH_SomMot_7 | RH_SomMot_33 |
| RH_SomMot_7 | RH_SomMot_34 |
| RH_SomMot_11 | RH_SomMot_32 |
| RH_SomMot_33 | RH_SomMot_34 |
| RH_SomMot_34 | RH_Amyg |

**Table S3**

List of connections between brain areas for the Metabolic Healthy/Obese (MHNW) > Metabolic unhealthy/Obese (MHO) connections.

| from | to |
| --- | --- |
| LH_SomMot_7 | Brainstem |
| LH_SomMot_11 | Brainstem |
| LH_SomMot_33 | RH_Cerebellum |
| LH_SalVentAttn_ParOper_1 | Brainstem |
| LH_Limbic_TempPole_1 | LH_Nacc |
| LH_Limbic_TempPole_1 | RH_Nacc |
| LH_Limbic_TempPole_3 | LH_Nacc |
| LH_Limbic_TempPole_3 | RH_Nacc |
| LH_Limbic_TempPole_6 | LH_Default_PFC_4 |
| LH_Limbic_TempPole_6 | LH_Default_PFC_9 |
| LH_Limbic_TempPole_6 | LH_Default_PFC_13 |
| LH_Limbic_TempPole_6 | RH_Default_PFCdPFCm_4 |
| LH_Limbic_TempPole_7 | LH_Default_PFC_1 |
| LH_Limbic_TempPole_7 | LH_Default_PFC_9 |
| LH_Limbic_TempPole_7 | RH_Cont_PFCmp_1 |
| LH_Limbic_TempPole_7 | RH_Default_PFCdPFCm_3 |
| LH_Limbic_TempPole_7 | RH_Default_PFCdPFCm_4 |
| LH_Default_PFC_4 | RH_Limbic_TempPole_2 |
| LH_Default_PFC_4 | RH_Default_Temp_1 |
| LH_Default_PFC_4 | Brainstem |
| LH_Default_PFC_4 | RH_Hipp |
| LH_Default_PFC_9 | RH_Default_Temp_1 |
| LH_Default_PFC_9 | Brainstem |
| LH_Default_PFC_12 | LH_Amyg |
| RH_Vis_11 | LH_Cerebellum |
| RH_Vis_11 | RH_Cerebellum |
| RH_SomMot_1 | Brainstem |
| RH_SomMot_3 | Brainstem |
| RH_SomMot_6 | Brainstem |
| RH_SomMot_7 | RH_Cont_PFCmp_2 |
| RH_SomMot_7 | Brainstem |
| RH_SomMot_9 | Brainstem |
| RH_SomMot_10 | Brainstem |
| RH_SomMot_11 | Brainstem |
| RH_SomMot_13 | Brainstem |
| RH_SomMot_14 | Brainstem |
| RH_SalVentAttn_TempOccPar_4 | Brainstem |
| RH_SalVentAttn_TempOccPar_5 | Brainstem |
| RH_SalVentAttn_TempOccPar_5 | LH_VentralDC |
| RH_SalVentAttn_PrC_1 | Brainstem |
| RH_SalVentAttn_FrOperIns_5 | RH_Nacc |
| RH_Limbic_OFC_3 | LH_Hipp |
| RH_Limbic_TempPole_1 | RH_Nacc |
| RH_Limbic_TempPole_4 | LH_Nacc |
| RH_Limbic_TempPole_6 | LH_Nacc |
| RH_Limbic_TempPole_6 | RH_Caudate |
| RH_Cont_PFCv_1 | RH_Nacc |
| RH_Cont_PFCmp_1 | RH_Default_Temp_3 |
| RH_Cont_PFCmp_1 | LH_Amyg |
| RH_Default_Temp_1 | RH_Default_PFCdPFCm_1 |
| RH_Default_Temp_1 | RH_Default_PFCdPFCm_3 |
| RH_Default_Temp_1 | RH_Default_PFCdPFCm_4 |
| RH_Default_PFCv_1 | Brainstem |
| RH_Default_PFCdPFCm_2 | RH_Caudate |
| RH_Default_PFCdPFCm_3 | LH_Amyg |
| RH_Default_PFCdPFCm_3 | Brainstem |
| RH_Default_PFCdPFCm_4 | Brainstem |
| RH_Default_PFCdPFCm_6 | LH_Amyg |
| LH_Nacc | Brainstem |
| LH_Nacc | LH_VentralDC |
| LH_Nacc | LH_Hipp |
| LH_Nacc | RH_Hipp |
| RH_Nacc | Brainstem |
| RH_Nacc | LH_Cerebellum |
| RH_Nacc | LH_VentralDC |
| RH_Nacc | RH_VentralDC |
| RH_Nacc | LH_Hipp |
| RH_Nacc | RH_Hipp |
| RH_Nacc | LH_Pallidum |
